# Developing a virtual Endoscopic Surgery Planning system to optimize surgical outcomes

**DOI:** 10.1101/2024.09.17.24313676

**Authors:** Bradley Hittle, Ahmad Odeh, Guillermo Maza, Brenda Shen, Bradley A Otto, Don Stredney, Gregory J. Wiet, Kai Zhao

## Abstract

**Objective:** Planning and predicting functional outcomes of endoscopic sinus surgeries (e.g., nasal airflow) based solely on visualizing Computerized Tomography (CT) or endoscopy poses a challenge to produce optimal clinical outcomes.

**Study design:** Technology development, retrospective case report.

**Methods:** A virtual surgery planning (VSP) tool is developed that can load any patient’s CT data and allow surgeons to remove obstructive tissue using both visual and haptic feedback endoscopically. Pre-calculated airflow resistance, wall shear stress, pressure drop are displayed on the anatomy to identify potential sites of obstruction. After each virtual surgery, changes in nasal airflow can be computed, and the process is reiterated until an optimal result is reached.

**Results:** As proof-of-concept, a series of isolated or combined procedures were performed on CT of one patient, who had olfactory losses that may involve obstructions blocking the air/odor flow to the olfactory fossa (OF). For this patient, an isolated medial partial middle turbinectomy (PMT) demonstrated the best outcome, better than traditionally performed lateral PMT, while septal body reduction worsened air/odor flow to OF.

**Conclusion:** This proof of concept case report demonstrates the potential usefulness of VSP in preoperative planning based on objective benchmarks and could be a valuable tool for optimizing future surgical outcomes.

## Introduction

Endoscopic nasal sinus surgery, pioneered by Messerklinger and further advanced by Stammberger and Kennedy^1^, has been the procedure of choice for the treatment of nasal obstruction and sinus disease, with an estimated 600,000 surgeries performed in the US every year^2^ alone. Nasal endoscopy provides landmark visualization and navigation for small instruments to operate within the nasal airway and sinuses. However, outcomes of these surgeries are highly variable, from short-term favorable outcomes of 60-90% to long-term outcomes as low as 30%^3,4^, potentially due to fact that planning and predicting functional outcomes (e.g., airflow) of endoscopic sinus surgeries can be difficult or deceptive based solely on visualizing Computerized Tomography (CT) or endoscopic images.

Virtual surgery planning (VSP) has been widely used in many other surgical fields that allows the surgeon to virtually plan the surgery to optimize surgical outcomes. Since previous endoscopic sinus surgery simulators^5–14^ mostly serve as surgical skill-training tools, we thus assembled an interdisciplinary team of engineers, computational scientists, and clinicians to develop an endoscopic sinus VSP system that utilizes state of the art computer graphics and 3D modeling that creates a desktop virtual environment to provide a prospective, preoperative guide that can optimize and improve surgical outcomes.

## Methods

The prototype comprises three distinct but synergistic components—a volume renderer to provide 3D endoscopic visualization based on patient’s clinical computerized tomography (CT) imaging; a haptic endoscopy tool to navigate and perform the virtual surgery; and the computational fluid dynamic (CFD) system to predict the nasal airflow outcome. The simulator runs on a desktop workstation, using an Intel Xeon E5-1650 v3 processor, 32GB of RAM, and an NVIDIA Quadro P5000 graphics card. The rendering system uses CUDA for Direct Volume Rendering (DVR) and OpenGL version 4.0 for graphics rendering that requires little to no preprocessing of CT data, no manual image segmentation, has fast frame rates and is extremely efficient.

A haptic device (PHANTOM, SensAble Technologies, Inc., Woburn, MA) that can apply forces in three degrees of freedoms, provides interaction between virtual endoscopic tools and the 3D CT-based volumetric model during virtual surgery. A virtual camera is attached to the endoscope tool, and the angle and distance of the tool tip can be adjusted, while the camera view is updated as the virtual endoscope navigates the nasal airway. The combination of 3D visual rendering and the haptic feedback allows the surgeon to remove the virtual obstructive tissue similar to in endoscopic surgery.

## Results

The developed virtual surgery planning (VSP) tool can load any patient’s CT in DICOM format in real-time from various sources, e.g. cone beam CTs or conventional axial spiral CTs, and provide a highly detailed 3D visual display of the nasal sinus airway (see Fig 1 and Supplementary method). The ability to directly load versatile DICOM-format clinical CT images without pre-processing is the key to allow this simulator to be utilized widely in clinical settings. Using a standard mouse input device, the user can orient the patient’s face/sinus anatomy in any direction. Pre-calculated airflow resistance, wall shear stress, pressure drop are displayed on the anatomy to identify potential sites of obstruction. In figure 1, a microdebrider was selected as the endoscopic tool, with available tool selections to be expanded in future software versions. The size of the microdebrider can be virtually adjusted. To ease operation, the surgical tool and camera are combined into a single haptic device, so that the surgeon can operate with one hand and toggle between different tools such as the endoscope and the microdebrider using buttons on the haptic device that are easily accessible by the user’s index finger. CT image guidance is also supported so that the surgeon can visualize the real-time tool tip location on 3 orthogonal CT slices on the side panels, mimicking the operating room (OR) experience^15^. Virtual surgeries can be saved intermittently (“mini save” button) or undone (“restore volume” button) to allow for easy planning with multiple surgical options. By restoring and saving multiple versions of different surgical approaches, different outcomes on a single patient can be compared and the process is reiterated until an optimal result is reached.

**Figure 1.**
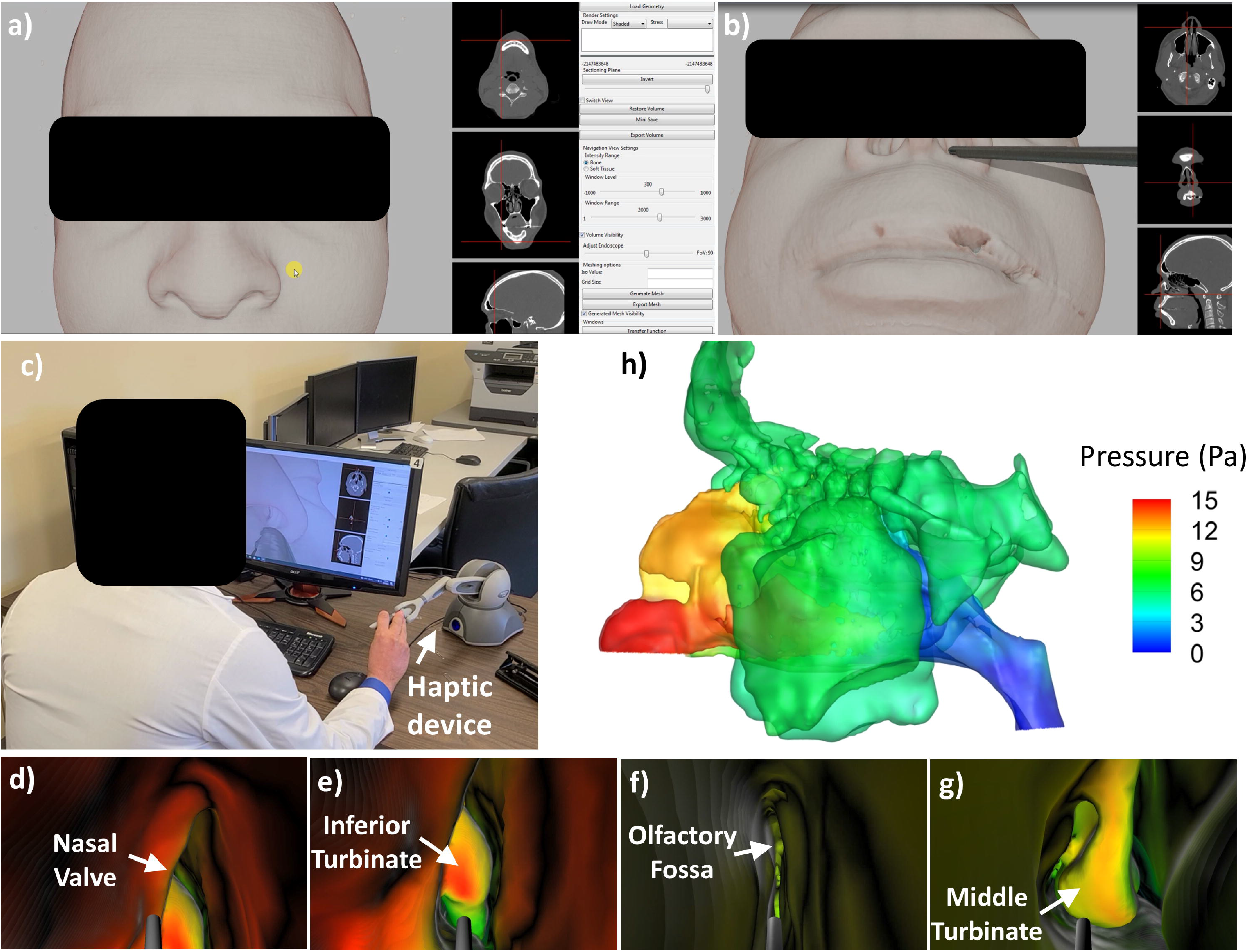
**(a) & (b)** Screen shots of nasal sinus VSP simulator with CT image guidance panels, shown with and without microdebrider selection **(c)** A surgeon operates the sinus surgery simulator. **(d-g)** Endoscopic view in the simulator at various anatomic landmarks within a patient’s nasal cavity. Colors indicate CFD-simulated air pressure during restful breathing **(h)**.

As proof-of-concept, we performed a series of isolated or combined procedures on one patient, who had nasal valve collapse, septal body hypertrophy, and a middle concha bullosa, and confirmed unilateral olfactory loss only to the left side (PEA threshold: left = 5, right = 9.3, normative range: >=8) that may involve obstructions blocking the air/odor flow to the olfactory fossa (OF). Before virtual surgery, this patient’s computed area-averaged PEA absorption flux to OF on the left side was significantly below the normal range (Fig 2e) that was established from 22 healthy controls collected from a prior study^16^, confirming the likely obstruction-driven olfactory loss, whereas the patient’s right pre-surgery olfactory PEA flux was within the normal range. Based on VSP, an isolated medial partial middle turbinectomy (PMT) demonstrated the best outcome, better than traditionally performed lateral PMT, while septal body reduction worsened air/odor flow to OF.

**Figure 2.**
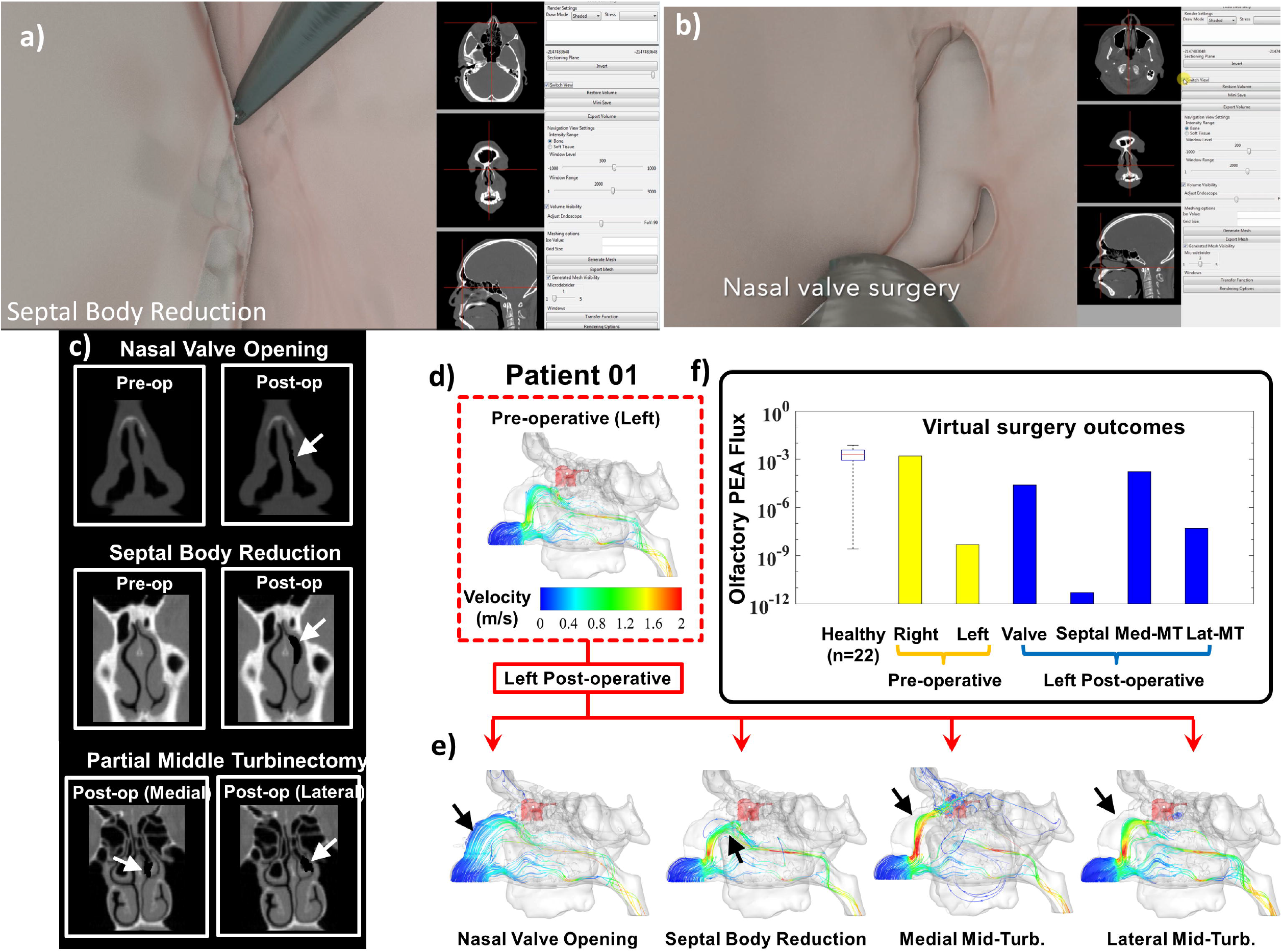
View of **(a)** septal body volume reduction (SBVR) and **(b)** nasal valvuloplasty virtual surgeries on patient #1 with unilateral nasal obstruction and olfactory loss (left side). **(c)** Pre- and post-virtual surgery CTs of various procedures. **Arrows** point to surgical sites. **(d, e)** Simulated **(d)** pre- and **(e)** post-virtual surgery airflow patterns in the left nasal airway. **Arrows** point to most significant air flow pattern changes. Among the various procedures performed, medial middle turbinate reduction and nasal valve expansion were more effective in directing airflow to the olfactory fossa (solid red shaded area). **(e)** Computed area-averaged phenyl ethyl alcohol (PEA: a rosy odor) absorption flux in the olfactory fossa pre- and post- virtual surgeries. An isolated medial aspect partial middle turbinectomy (Medial Mid-Turb) showed the best outcome, and it was better than traditionally performed lateral Mid-Turb, while septal body reduction worsened air/odor flow to OF. Surprisingly, surgery to the nasal valve also leads to good outcome. Before virtual surgery, the left olfactory PEA absorption flux is significantly below that of healthy controls (n=22), confirming the obstruction-related loss.

## Discussion

This proof of concept trial demonstrates the potential usefulness of VSP in preoperative planning based on objective benchmarks and could be a valuable tool for optimizing future surgical outcomes. In contrast to previous systems which emphasize training^5–14^, the current VSP emphasizes on surgical planning. The endoscope environment and haptic feedback only serve as an intuitive interface for experienced surgeons to remove obstructive tissue and plan for surgery in a familiar and more effective way than say on CT scan or using a keyboard and mouse. The integration of an intuitive interface, ability to easily load patient image data, with clinically relevant outcome data (CFD) are significant advancements over previous attempts of sinus surgical planning simulators ^17^. Such a well-implemented planning tool is likely to have a higher transfer rate to the OR for practicing surgeons and broader clinical impact than those only for surgical training purposes. Finally, the optimized surgical plans can potentially be translated into a CT scan (see Fig 1) and loaded into image guidance systems in ORs to guide the surgeon during surgery, which may complete the full circle of personalized medicine: from planning, optimizing, and to the operating room.

## Data Availability

All data, code, and materials used in the analysis are available to any researcher for purposes of reproducing or extending the analysis via institutional materials transfer agreements (MTAs).

## Acknowledgements

We acknowledge Bhakthi Deshpande, Chengyu Li and Zhenxing Wu of the Department of Otolaryngology - Head & Neck Surgery, the Ohio State University, for their assistance in patient testing as well as in CFD simulations.

## Author contributions

Conceptualization, Methodology, Project administration, Supervision, Funding acquisition: KZ, GJW, DS

Investigation: BH,GM, AO, BS, BAO, DS, GJW, KZ

Visualization: GM, BH, AO, BS, KZ

Writing – original draft: KZ

Writing – review & editing: KZ, AO, GM, BH, GJW

## References

1. Kennedy DW, Zinreich SJ, Rosenbaum AE, Johns ME. Functional endoscopic sinus surgery. Theory and diagnostic evaluation. Arch Otolaryngol Chic Ill 1960. 1985;111(9):576–582. doi:10.1001/archotol.1985.00800110054002

2. Bhattacharyya N. Ambulatory sinus and nasal surgery in the United States: demographics and perioperative outcomes. The Laryngoscope. 2010;120(3):635–638. doi:10.1002/lary.20777

3. Stewart MG, Smith TL, Weaver EM, et al. Outcomes after nasal septoplasty: results from the Nasal Obstruction Septoplasty Effectiveness (NOSE) study. Otolaryngol--Head Neck Surg Off J Am Acad Otolaryngol-Head Neck Surg. 2004;130(3):283–290. doi:10.1016/j.otohns.2003.12.004

4. Ho W kuen, Yuen APW, Tang K chi, Wei WI, Lam PKY. Time course in the relief of nasal blockage after septal and turbinate surgery: a prospective study. Arch Otolaryngol Head Neck Surg. 2004;130(3):324–328. doi:10.1001/archotol.130.3.324

5. Edmond CV, Heskamp D, Sluis D, et al. ENT endoscopic surgical training simulator. Stud Health Technol Inform. 1997;39:518–528.

6. Rudman DT, Stredney D, Sessanna D, et al. Functional endoscopic sinus surgery training simulator. The Laryngoscope. 1998;108(11 Pt 1):1643–1647. doi:10.1097/00005537-199811000-00010

7. Hilbert M, Müller W, Strutz J. [Development of a surgical simulator for interventions of the paranasal sinuses. Technical principles and initial prototype]. Laryngorhinootologie. 1998;77(3):153–156. doi:10.1055/s-2007-996951

8. Nash R, Sykes R, Majithia A, Arora A, Singh A, Khemani S. Objective assessment of learning curves for the Voxel-Man TempoSurg temporal bone surgery computer simulator. J Laryngol Otol. 2012;126(7):663–669. doi:10.1017/S0022215112000734

9. Varshney R, Frenkiel S, Nguyen LHP, et al. The McGill simulator for endoscopic sinus surgery (MSESS): a validation study. J Otolaryngol - Head Neck Surg J Oto-Rhino-Laryngol Chir Cervico-Faciale. 2014;43(1):40. doi:10.1186/s40463-014-0040-8

10. Won TB, Hwang P, Lim JH, et al. Early experience with a patient-specific virtual surgical simulation for rehearsal of endoscopic skull-base surgery. Int Forum Allergy Rhinol. 2018;8(1):54–63. doi:10.1002/alr.22037

11. Lindquist NR, Leach M, Simpson MC, Antisdel JL. Evaluating Simulator-Based Teaching Methods for Endoscopic Sinus Surgery. Ear Nose Throat J. 2019;98(8):490–495. doi:10.1177/0145561319844742

12. Kim DH, Kim HM, Park JS, Kim SW. Virtual Reality Haptic Simulator for Endoscopic Sinus and Skull Base Surgeries. J Craniofac Surg. 2020;31(6):1811–1814. doi:10.1097/SCS.0000000000006395

13. Richards JP, Done AJ, Barber SR, Jain S, Son YJ, Chang EH. Virtual coach: the next tool in functional endoscopic sinus surgery education. Int Forum Allergy Rhinol. 2020;10(1):97–102. doi:10.1002/alr.22452

14. Carsuzaa F, Fieux M, Legre M, et al. SimLifeÂ®, a new dynamic model for endoscopic sinus and skull base surgery simulation. Rhinology. 2023;61(6):574–576. doi:10.4193/Rhin23.230

15. Fried MP, Morrison PR. Computer-augmented endoscopic sinus surgery. Otolaryngol Clin North Am. 1998;31(2):331–340. doi:10.1016/s0030-6665(05)70052-9

16. Li C, Jiang J, Kim K, et al. Nasal Structural and Aerodynamic Features That May Benefit Normal Olfactory Sensitivity. Chem Senses. 2018;43(4):229–237. doi:10.1093/chemse/bjy013

17. Burgos MA, Sevilla García MA, Sanmiguel Rojas E, et al. Virtual surgery for patients with nasal obstruction: Use of computational fluid dynamics (MeComLand®, Digbody® & Noseland®) to document objective flow parameters and optimise surgical results. Acta Otorrinolaringol Esp. 2018;69(3):125–133. doi:10.1016/j.otorri.2017.05.005

