## Supplemental Method for "Developing a virtual Endoscopic Surgery Planning system to optimize surgical outcomes"

**Supplementary Methods**

The VSP uses a scene to manage all information within the simulator that includes CT data, lights, camera and tools. To increase the fidelity and physical accuracy of the simulation in the future, the renderer of the scene is a hybrid rasterizer and ray tracer using physically based rendering techniques shown by Karis ^1^ and Kroes et al.^2^ that occurs in three steps: tool rendering, CT volume rendering, and finally compositing. The tool is represented by triangular meshes that uses physically based material properties for lighting that contributes to the realism of the scene. These triangular meshes are rendered with OpenGL 4.0 specification (<https://registry.khronos.org/OpenGL-Refpages/gl4/>) through rasterization into a geometric buffer (g-buffer)^3^ using Image-Based Lighting (IBL).^4^ CUDA 8.0 performs the Direct Volume Rendering (DVR) using ray casting to visualize CT data. The DVR was designed to render the CT data as fast as possible without the need for pre-processing. The DVR will draw the bounding box in normalized device coordinates in an offscreen buffer, which is used as a setup step before performing any ray casting. The DVR will then read the CT bounding box and use this information to determine ray directions. For every point in the camera’s view, a ray is released and marched, sampling intensity points in the CT volume along the way, until it exits the CT bounding box or hits a recorded g-buffer object.  These samples are converted into colors and opacities using a transfer function and output into a secondary buffer. Once the ray marching buffer has been completed, a screen-space composite shader determines how each pixel within the camera view will be drawn for the scene. The final step composites the two buffers into a cohesive scene which is displayed to the user’s screen. This is necessary for compositing objects within the transparent volumetric data in the scene. The provided g-buffers allow for shadows, ambient occlusion, and motion-blur to increase the realism of the scene. The overall design caches each drawn image and supports regional rendering to re-draw only regions that have been modified.

For haptic feedback, we use a physically motivated model designed by Sonny Chan at the University of Calgary.^5^  It uses a 6-degree-of-freedom, constraint-based technique that employs a proxy device to keep the cutting instrument in a penetration-free state. This technique enforces a non-intersection condition between the simulated instrument and the nasal anatomy which better reflects the situation and makes it robust to instabilities.  Being physically based, the amount of removal is directly tied to simulated physical properties of the material, cutting geometry, and the amount of force applied by the user. While the haptics thread updates at a frequency of around 2,000 Hz, removal only occurs at a rate of 200 Hz.  The default removal uses the Archard’s derivation^6^ with a radius of 0.5mm. Each removal tick solves the equation:

∆V = k F_n_ ∆x / 3H	(s1)

Where F_n_ is the applied load or normal force, ∆V is the quantity of volume to remove, ∆x is the distance between the two surface contacts, H indicates the Vickers hardness, and finally, k is a dimensionless wear coefficient. Since the prototype developed this study is not intend to replicate real endoscopic surgery (such as that used for training), rather the endoscope environment and haptic device are only intend to allow surgeons to remove obstructive tissue and plan for surgery in a familiar and more effective way than say directly on CT scan or using a keyboard and mouse, thus, H is set to the hardness value of bone according to literature^7^ and k is set to =1, which represents completely removal of the tissue once contacts the cut surface, regardless of tissue biomechanical properties. The cut surface is a cylinder that outlines the debrider tool tip, which has a blade and a non-blade side. An additional algorithm limits the removal only on the blade side of the debrider, while allowing the tip to be against tissue on the non-blade side. Once ∆V is calculated, removal of material from neighboring voxels occurs by shelling the proxy outward.

The VSP software was developed using C++ 14 and compiled using Microsoft Visual Studio 2017. The user interface was created using wxWidgets 3.12 (<https://www.wxwidgets.org/>). The OpenGL library Glew (<https://glew.sourceforge.net>) was used for loading extensions. The mathematics library GLM (<https://github.com/g-truc/glm>) was used for calculations . The OpenHaptics SDK (<https://www.3dsystems.com/haptics-devices/openhaptics>) is used to communicate with the Sensable PHANToM device. Eigen3 (<https://eigen.tuxfamily.org/index.php?title=Main_Page>) is used for numerical solutions. CT Data is loaded using GDCM (<https://sourceforge.net/projects/gdcm/>) and is written with Nifti (<https://github.com/NIFTI-Imaging/nifti_clib>). The environment map is loaded with GLI (<https://github.com/g-truc/gli>). The triangular meshes are loaded using the asset importer library (<https://www.assimp.org/>). Intel’s oneAPI Thread Building-blocks (https://github.com/oneapi-src/oneTBB) is used for multi-threading.

Reference:

1.	Karis, B. SIGGRAPH 2013 Course: Physically Based Shading in Theory and Practice. Published 2013. Accessed November 22, 2023. https://blog.selfshadow.com/publications/s2013-shading-course/

2.	Kroes, P.F, Botha CP. Correction: Exposure Render: An Interactive Photo-Realistic Volume Rendering Framework. *PLoS ONE*. 2013;8(4).

3.	Saito T, Takahashi T. Comprehensible rendering of 3-D shapes. *SIGGRAPH Comput Graph*. 1990;24(4):197-206. doi:10.1145/97880.97901

4.	Debevec P. Image-based lighting. In: *ACM SIGGRAPH 2006 Courses on	- SIGGRAPH ’06*. ACM Press; 2006:4. doi:10.1145/1185657.1185686

5.	Chan S, Conti F, Blevins NH, Salisbury K. Constraint-based six degree-of-freedom haptic rendering of volume-embedded isosurfaces. In: *2011 IEEE World Haptics Conference*. IEEE; 2011:89-94. doi:10.1109/WHC.2011.5945467

6.	Archard, J.F. Contact and Rubbing of Flat Surfaces. *Journal of Applied Physics*. 1953;24(8):981-988.

7.	Zysset PK, Guo XE, Hoffler CE, Moore KE, Goldstein SA. Elastic modulus and hardness of cortical and trabecular bone lamellae measured by nanoindentation in the human femur. *J Biomech*. 1999;32(10):1005-1012. doi:10.1016/s0021-9290(99)00111-6

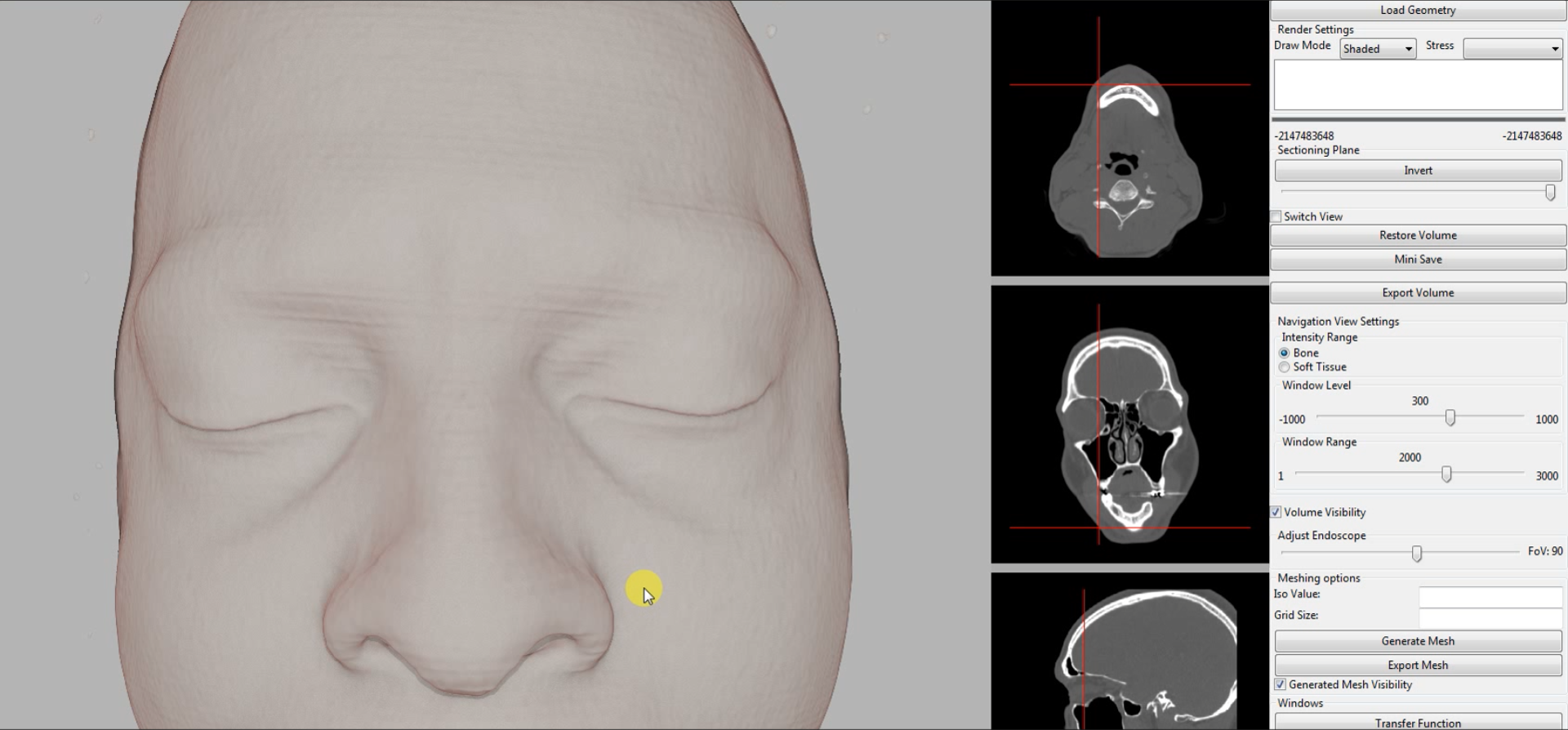

Supplementary Figure 1.

User interface of the surgical simulator.
